# Fatigue outcomes following coronavirus or influenza virus infection: A systematic review and meta-analysis

**DOI:** 10.1101/2020.12.04.20244145

**Authors:** Kim Poole-Wright, Fiona Gaughran, Rachael Evans, Trudie Chalder

## Abstract

**Objectives:** Fatigue is a pervasive clinical symptom for many infected with respiratory viruses such as influenza or coronaviruses. Prior evidence from influenza and coronavirus epidemics suggest that fatigue symptomology may continue beyond the acute phase, lasting for several months to several years post-discharge. This systematic review aimed to examine long-term fatigue prevalence among survivors and among communities, as well as investigate the current evidence for associated factors.

**Design:** Systematic review and meta-analysis.

**Setting:** Hospitalised and community samples.

**Participants:** Patient populations with a confirmed diagnosis of a named influenza virus or coronavirus.

**Main outcomes measured:** Fatigue, fatigue syndromes

**Results:** Ten studies met the inclusion criteria for a pooled prevalence analysis and five studies were identified as eligible for a means differences analysis. A fatigue prevalence of 41% (95% CI 0.299-0.488) was found among a total population of 1,310. Using the ‘vitality’ subscale of the SF-36 as a proxy for fatigue, the estimate for means differences indicated a lower mean vitality score for survivors compared to population norms (M -1.523, CI -13.53 – 10.48), although this was not significant (p = 0.803). The most common associations with fatigue were PTSD, depression and anxiety, female gender and higher age.

**Conclusions:** This study reveals that a significant proportion of survivors (41%) experienced fatigue following their recovery from novel respiratory viruses such as SARS, MERS, SARS-CoV2 or influenza and that this fatigue can be long-lasting. Also, some factors such as female gender and psychological factors may contribute to continuing fatigue outcomes for this population.

**Strengths and limitations:** (a) this study provides support for long-term fatigue outcomes in people with a confirmed influenza, SARS, MERS, SARS-CoV2 virus infection (b) the study suggests individual, psychological and social factors are associated with fatigue, (c) findings are limited by the availability of fatigue data and lack of pre-morbid fatigue information; (d) a meta-analysis on the associations was prohibited by the small number of studies investigating long-term fatigue correlates and (e) the heterogeneity of the studies (>75%) suggests the pooled estimates should be interpreted with caution.

## INTRODUCTION

Coronavirus family such as SARS-CoV Severe Acute Respiratory Syndrome (SARS), Middle East Respiratory Syndrome (MERS) and SARS-CoV-2 are single-strand RNA viruses, characterised by severe upper respiratory tract infections. Influenza viruses can also result in severe respiratory symptoms. A novel A(H1N1)pdm09 influenza emerged in humans in 2009 and before this other strains, originating in animals or birds had circulated, including the influenza virus A subtype H2N2 and HPAI A(H5N1). Each has spread rapidly worldwide with considerable impact on mortality. The case fatality rate for SARS during the course of the outbreak (2002 - 2004) was 9.7% (Petersen et al., 2020). For MERS an estimated 2,494 cases were detected since its identification in 2012, with a fatality rate of 34.4% (Bradley & Bryan, 2019). H3N2 mortality ranged from 0.03% and over 18,400 worldwide deaths were attributed to H1N1 (WHO, 2015). H2N2 had a fatality rate of 1.9/10,000 population (Viboud et al., 2015). Approximately 861 laboratory-confirmed cases of A(H5N1) have been reported (WHO, 2020) and for SARS-CoV2 current worldwide estimates are reported to be 67,965,261 cases (ECDC, 2020). Wide-ranging clinical indicators have been observed for some infectious viruses from asymptomatic (Leung, Xu, Ip, & Cowling, 2015) to the most severe, potentially resulting in respiratory distress or viral pneumonia. Outcomes are dependent upon variables such as age and physical comorbidities. Fatigue is among the most common presenting symptom alongside high fever, myalgia, cough and breathlessness. Clinical observations for patients infected with viruses such as SARS have indicated fatigue in 78% of patients admitted to hospital (Han et al., 2003). Meta-analyses of the current SARS-CoV-2 (COVID-19) virus indicates prevalence to be between 22-61.9% (Qiu et al., 2020; Wang et al., 2020) and fatigue has been found to be associated with exacerbated outcomes, including acute respiratory distress and mortality. A study investigating COVID-19 found that, compared with survivors, more severe fatigue was associated with mortality in a small number of patients admitted to an ICU (Wang, Shu, Ran, Xie, & Zhang, 2020).

Fatigue may be characterised as tiredness or exhaustion as a result of physical or mental exertion or as a result of an illness or disease (Dittner, Wessely, & Brown, 2004). The experience of fatigue is common and is usually short-lived but, for a small number of people, it can become long-lasting, associated with a number of impairments in daily living and reduced quality of life (Dittner et al., 2004).

Post-viral fatigue has been linked to viruses including Epstein-Barr and Q Fever, while chronic fatigue has been investigated in survivors of SARS, MERS and H1N1. Chronic fatigue syndrome has been associated with the H1N1 virus for instance (Magnus et al., 2015) and, in 233 SARS survivors, 43% had chronic fatigue, with around 27% fulfilling the criteria for chronic fatigue syndrome 3 years post-infection (Lam et al., 2009). A meta-analysis comparing SARS, MERS and COVID-19 found a post-illness fatigue prevalence of 19.3% (Rogers et al., 2020). Other research indicates the development of a post-SARS syndrome for some survivors (Chrousos & Kaltsas, 2005; Moldofsky & Patcai, 2011). There is emerging evidence for lasting fatigue as a consequence of COVID-19. A recent study found moderate to severe levels of fatigue in up 54% of 100 patients treated for coronavirus, 4 weeks post-discharge (Halpin et al., 2020) and at a median follow-up stage of 10 weeks, up to 50% reported severe fatigue (Townsend et al., 2020).

Factors affecting post-illness fatigue have been investigated in previous viral influenza outbreaks. Studies indicate that for many SARS survivors, lung function was well preserved with significant improvements between 3-12 months post-infection, although remaining lower than healthy controls (Hui et al., 2005; Ong, 2004). Similar results have been found for exercise capacity. For example, several studies have found poorer exercise performance on the six-minute walking test (6MWT) at 3, 6 and 12 months (Hui, 2005; Ong, 2004). These exercise impairments, however, were not correlated with lung function. For example, in a Singapore study of 44 SARS patients, 41% had reduced exercise capacity but mild pulmonary function defects (Ong, 2004). These findings have been replicated in other SARS survivors (Hui et al., 2009; Hui et al., 2005; Ngai et al., 2010; Tansey et al., 2007) and H1N1 patients (Hsieh et al., 2018). These physical functioning limitations, independent of pulmonary function are thought to contribute to the reduced quality of life and fatigue outcomes found in numerous studies of recovered patients (Hsieh et al., 2018; Tansey et al., 2007)

These results suggest that, alongside possible physical deconditioning and corticosteroid myopathy (Dekhuijzen & Decramer, 1992; Tansey et al., 2007), other psychological, social or cognitive contributions may explain the persistence of fatigue and lower QoL, for example, long periods of social isolation or stigma (Wing & Leung, 2012). Time spent in ICU (Halpin et al., 2020) and hospital length of stay (Chen et al., 2020) have been associated with fatigue for instance and psychiatric disorders such as anxiety, post-traumatic stress disorder and depression are high in post-SARS. Further, chronic fatigue has been associated with a risk of psychiatric disorder in recovered SARS patients (Wing & Leung, 2012) and with post-traumatic stress symptoms in MERS survivors (Lee et al., 2019). Social factors have been found to be a factor in post-discharge fatigue. For instance, fatigue was more likely where patients were involved in ‘medicolegal’ cases (Wing & Leung, 2012) and, controlling for psychiatric morbidity, being an applicant to the survivors’ fund in Hong Kong (Lam et al., 2009). Furthermore, while around 76-80% of survivors achieve a return to work (Lau et al., 2005; Rogers et al., 2020), some patients reported fatigue as one of the reasons for not returning to work (Guo et al., 2019).

### Summary

Fatigue for the majority of patients usually dissipates during the course of a virus, but some experience longer lasting symptoms, independent of their pulmonary functions and exercise capacity. These symptoms may be associated with a number of psychosocial factors such as psychiatric morbidity and isolation, which have been indicated with fatigue. Long-term consequences of an epidemic such as SARS, MERS or H1N1 revealed by prior research, may be pertinent to survivors of the current COVID-19 pandemic. Therefore, the aim of this systematic review was to investigate the prevalence of persistent fatigue among survivors of a viral epidemic and consolidate current evidence for factors associated with fatigue outcomes.

## METHODS

### Search strategy

The protocol and PICO framework for this study was developed utilising The Preferred Reporting Items for Systematic Reviews and Meta-Analyses (PRISMA). The databases Embase, PsyINFO, Medline, CINAHL, Cochrane Database of Systematic Reviews and Open Grey were searched from inception dates to (open date), using defined search terms: H1NI OR “swine flu” OR “swine influenza OR SARS OR “severe acute respiratory syndrome” OR coronavirus OR COVID-19 OR H5NI OR “avian flu” OR “avian influenza” OR MERS OR “Middle East respiratory syndrome” OR H3N2 OR “Hong Kong flu” OR “Hong Kong influenza” OR H2N2 OR “Asian flu” OR “Asian influenza” OR pandemic OR epidemic AND “chronic fatigue” OR fatigue OR exhaustion OR “quality of life” OR HRQoL. MeSH (Medical Subject Headings) were included in order to maximise the search. Reference lists of the selected studies were manually searched for additional articles.

Eligibility criteria

- Studies were considered eligible for inclusion if they met the following criteria:
  - Original articles available in English;
  - Studies with primary data;
  - Studies reporting fatigue or fatigue syndromes using a fatigue measure or a fatigue/tiredness related subscale;
  - Studies investigating fatigue occurring post-discharge;
  - Studies investigating patient populations diagnosed with either COVID-19, severe acute respiratory syndrome (SARS), middle east respiratory syndrome (MERS), influenza A/H1N1pdm09 (Swine), HPAI H5N1 (Avian), H2N2 (Asian), A/H3N2 (Hong Kong) or other sub-virus;
  - Any study design including cohort, case-control, cross-sectional, randomised control trials, meta-analysis.

Studies were excluded if they (1) measured compassion or pandemic fatigue (defined as ‘worn out’ by pandemic warnings, by government safety instructions, or with media coverage, or with compliance requirements; (2) populations without a confirmed diagnosis of one of the named viruses; (3) reported fatigue associated with physical disorders (e.g. thyroiditis, Parkinson’s disease, cancer); (4) measured fatigue as a clinical symptom during the acute phase (defined as the period of hospitalisation); (5) were protocols, vaccination studies, animals; (6) did not report fatigue outcome data; (7) were newspaper articles, conference papers, commentaries or editorials. We also excluded qualitative studies and studies investigating fatigue among healthcare workers.

### Data extraction

Titles and abstracts were screened by the first researcher (KPW). Full texts were screened by KPW against the eligibility criteria. A data spreadsheet was created to include extracted data from the included studies. The senior researchers (TC, FG) reviewed a number of the final included studies and data. PRISMA Flow Diagram available in Appendix A. Discrepancies were resolved via discussion and consensus with TC. The variables obtained for the data synthesis spreadsheet were: citation, population, number of participants, control group, location, virus type, follow-up period, study design, outcome variable of interest (e.g. fatigue, vitality), associated variables (e.g. PTSD, depression, anxiety, stress), scales or measures employed, power calculation (Y/N).

### Quality Assessments

Risk of bias was assessed by the Critical Appraisal Skills Programme (CASP) (2018). Each study design has an appropriate checklist (e.g. cohort) comprising 12 items designed to systematically assess a study. The items do not require a score but rather a “yes”, “no” or “can’t tell”. The randomised and systematic review checklists were developed and adapted from JAMA’s guides to medical literature. An overall assessment was made by assigning a grade of ‘excellent’, ‘good’, ‘moderate’ or ‘low’ to each included article.

### Statistical analysis

We computed pooled prevalence for binary fatigue outcomes with 95% confidence intervals for 10 studies. A number of studies investigated fatigue outcomes across multiple time points. Therefore, in order to maintain the independence of observations, only one time period was included in the analysis as follows: (a) studies investigating fatigue in the weeks following an infection, we selected the period closest to 3 months and (b) studies with 12 months or longer time periods, we selected the period closest to 12 months. This was to demonstrate that the fatigue assessed was ‘long-term’. Where studies investigated both ‘fatigue’ and CFS outcomes, we incorporated the fatigue data only in the meta-analysis. This was because the participants had not received a confirmed diagnosis of CFS in all studies. Additional meta-analysis was conducted to compute mean differences from five included studies to estimate the ‘severity’ of fatigue compared to population norms. For this computation, the ‘Vitality’ subscale of the Medical Outcomes Study 36-item Short-Form health survey (SF-36) (Ware & Sherbourne, 1992) was utilised as a proxy for fatigue. This ‘vitality’ subscale is scored from 0-100, with higher scores representing better functioning/less disability. For the purposes of this study lower scores represent lower ‘energy’ and higher fatigue. We utilised this instrument because it the most frequently used measure assessing HRQoL in respiratory virus research. The absence of data from control groups precluded other analyses except population norms. Effect sizes were first calculated from each included study to compute the mean effect. Meta-analyses were conducted using R Studio, Version 1.3.1073 (2020). Heterogeneity was assessed using Cochran Q statistic. We obtained the 1^2^ statistic with the degree of heterogeneity categorised as ‘not important’ (0-40%), ‘moderate’ (30-60%), ‘substantial’ (50-90%) and ‘considerable’ (75-100%) (Higgins, 2003). We conducted an Egger’s test to explore potential publication bias for our proportional analysis: bias = 1.657 (95% CI -7.19-10.5), p = 0.723 suggesting an absence of funnel plot asymmetry. Due to the small number of studies further analysis of the origins of heterogeneity via a meta-regression or the production of funnel plots was not performed on the mean difference analysis.

Multiple studies investigated a wide-range of post-virus outcomes including psychological, physical and/or social effects. A number of these investigated the relationship between an exposure virus and fatigue outcomes. However, each used a diverse number of outcome measures or utilised non-validated scales, which disqualified the viability of a meta-analysis. Consequently, associations which had available data were arranged in tabular form illustrating the direction of the association with fatigue or vitality (Table 2). A positive symbol (+) indicated a positive association, a negative symbol (-) indicated a negative association and a zero (0) indicated no significant association between the investigated variable and fatigue (Matcham, Ali, Hotopf, & Chalder, 2015). Associations with fatigue or vitality, measured in prospective cohort designs were demonstrated by a superscript figure representing the period the relationships were examined.

## RESULTS

### Search results

A total of 2,557 articles were identified using the database search protocols. Following the removal of duplicates, 2,108 articles remained for title and abstract screening. Of these a total of 201 articles were selected for full text screening producing a final total of 32 studies meeting the eligibility criteria and 15 were deemed eligible for a quantitative analysis. A summary of the included articles is presented in Table 1. We tabulated the studies according to the fatigue and ‘vitality’ outcome measures.

**Table 1.**
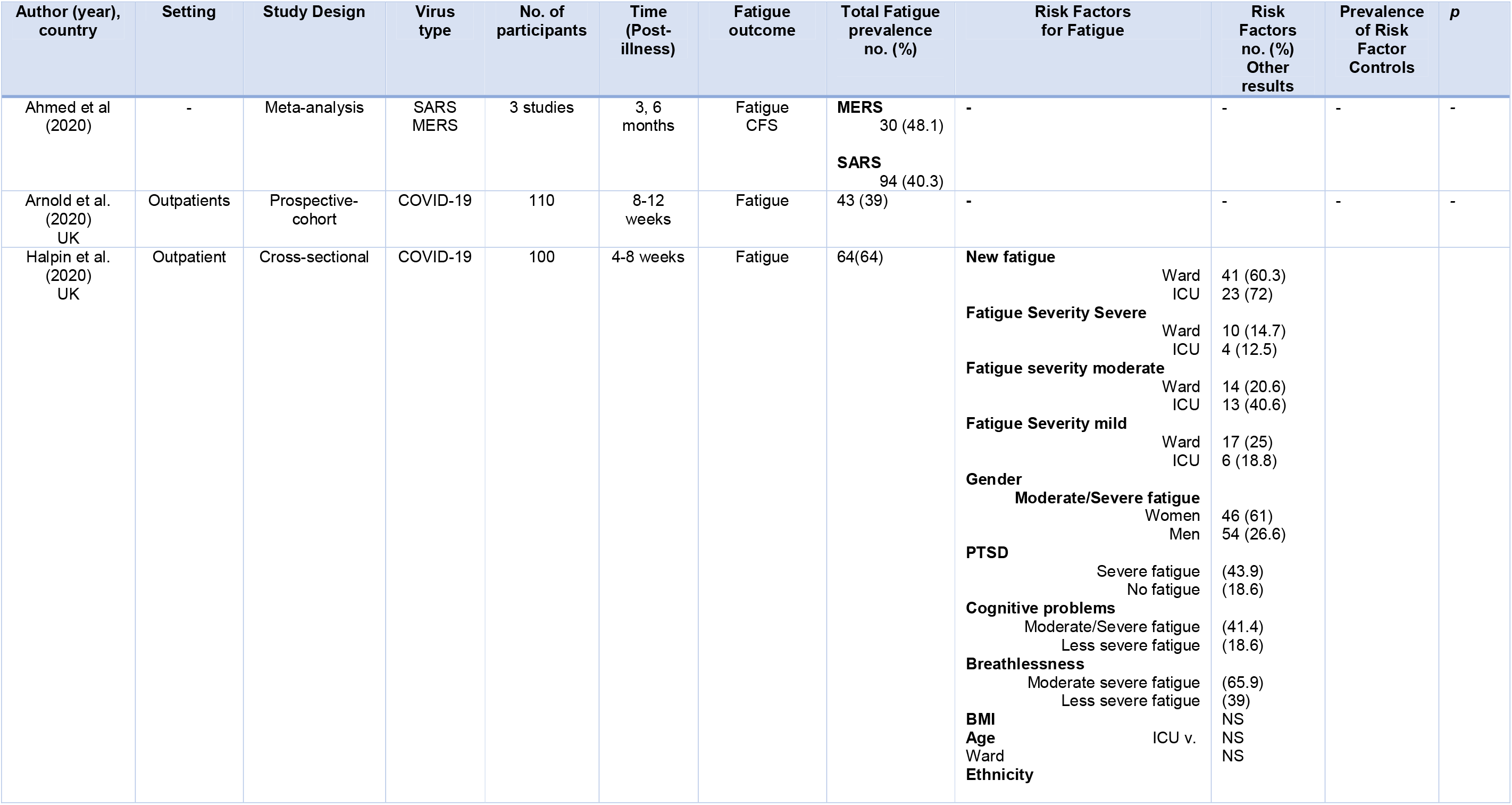
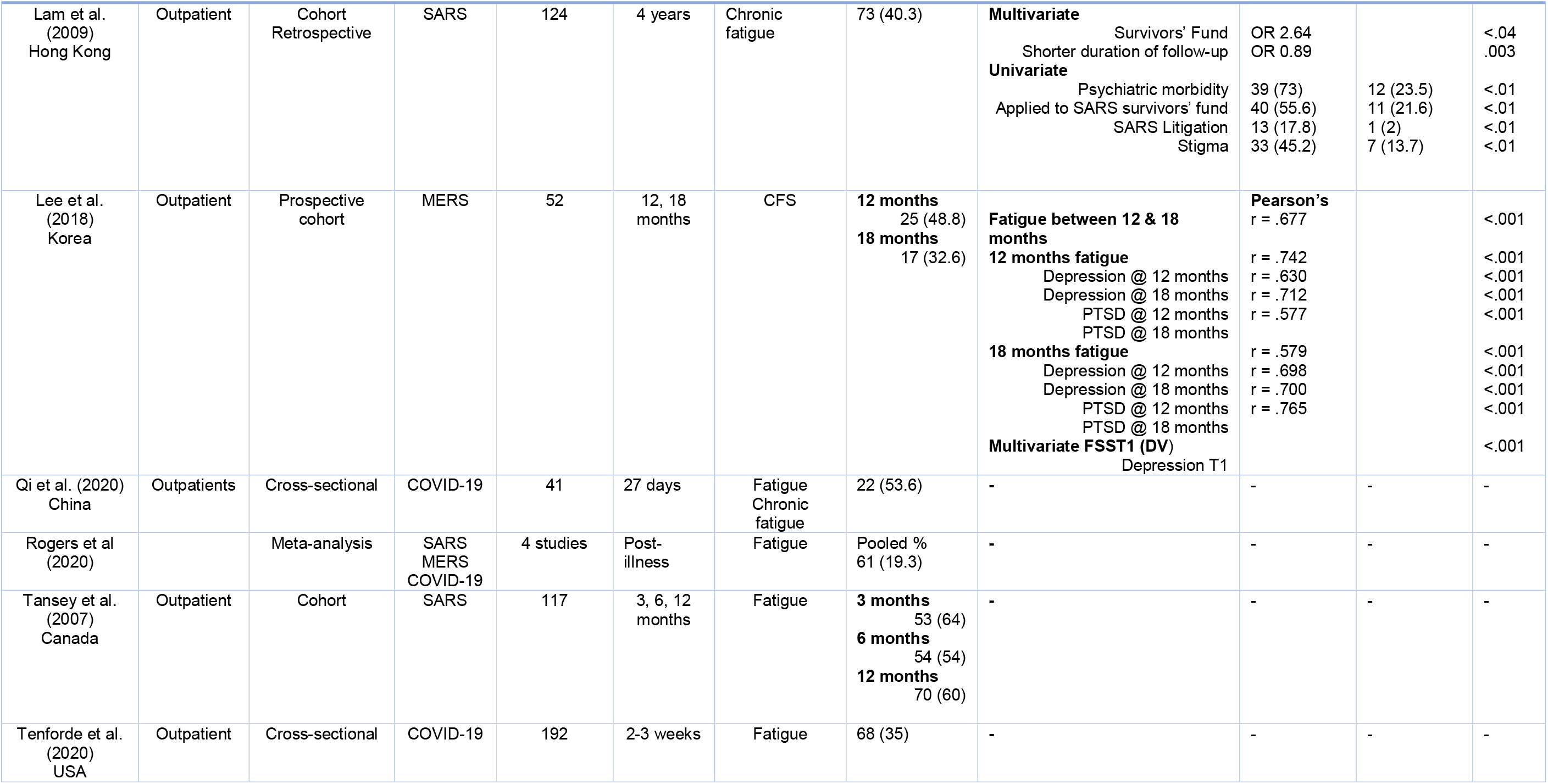
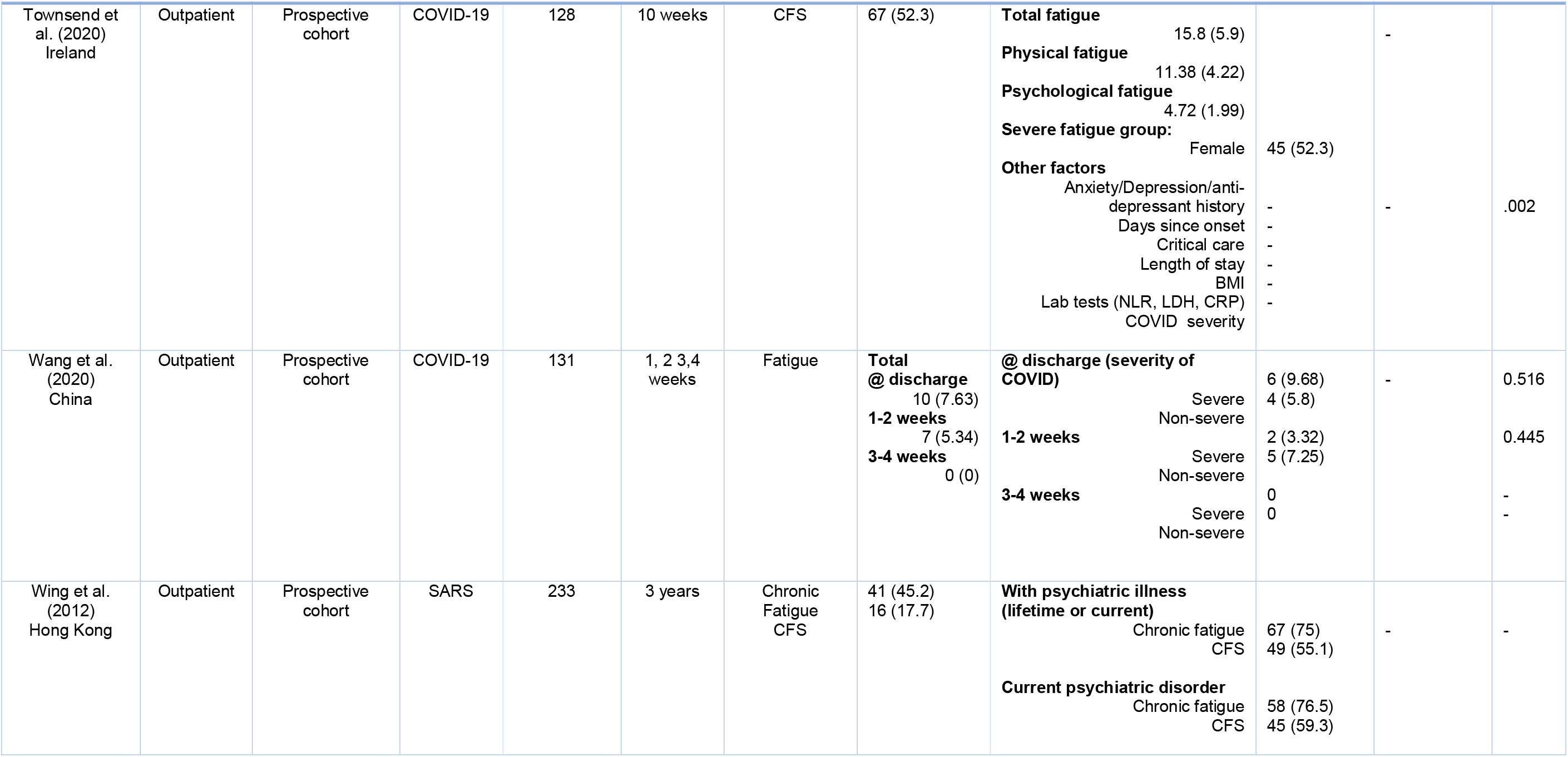
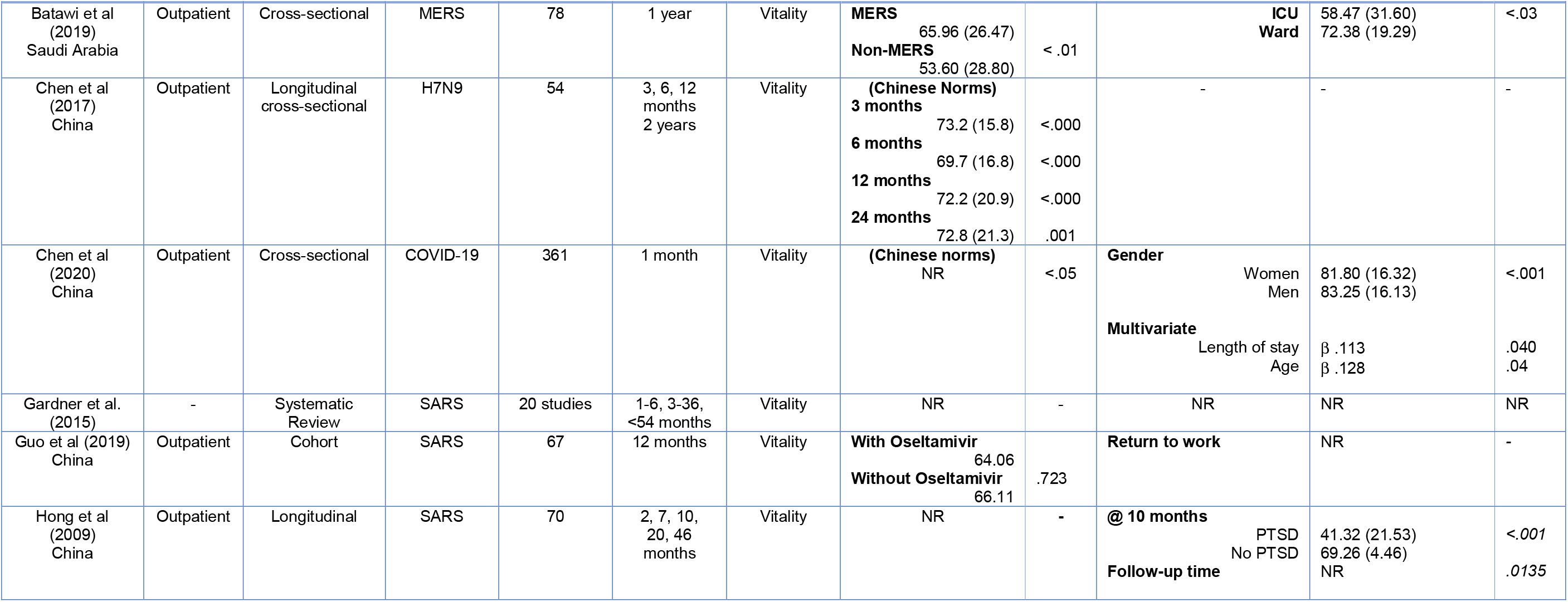
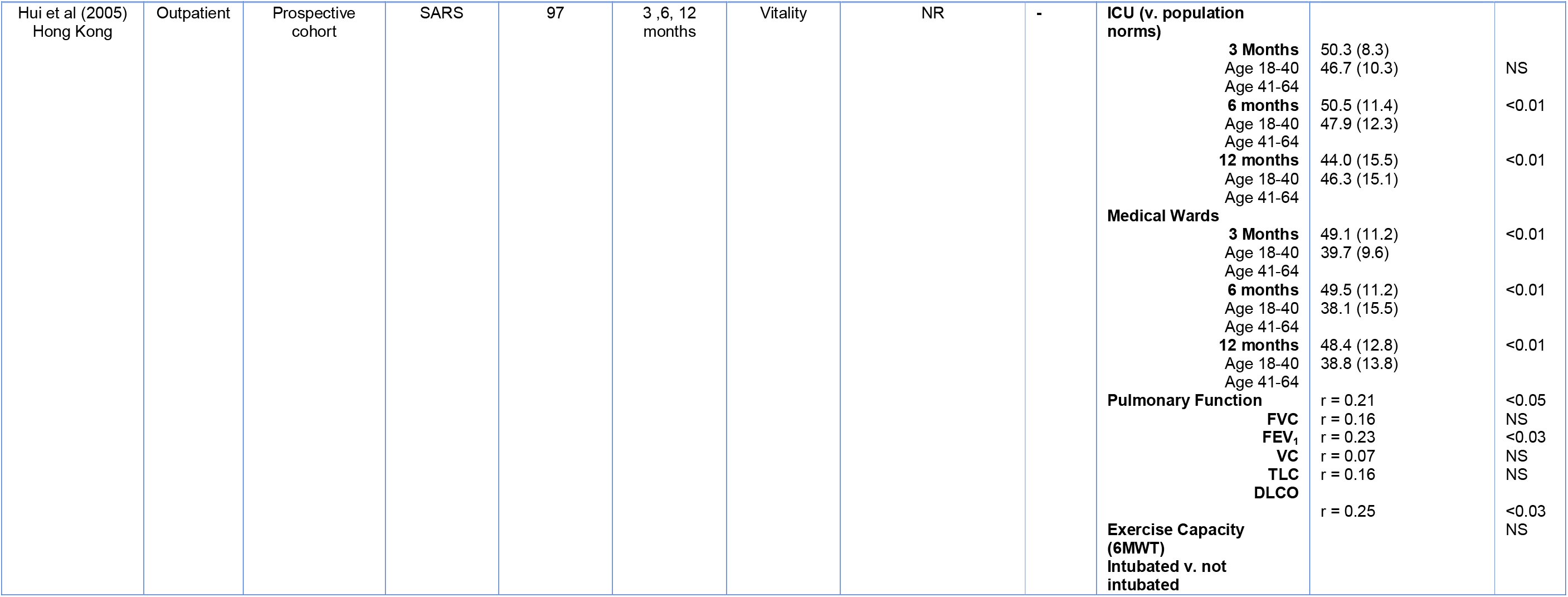
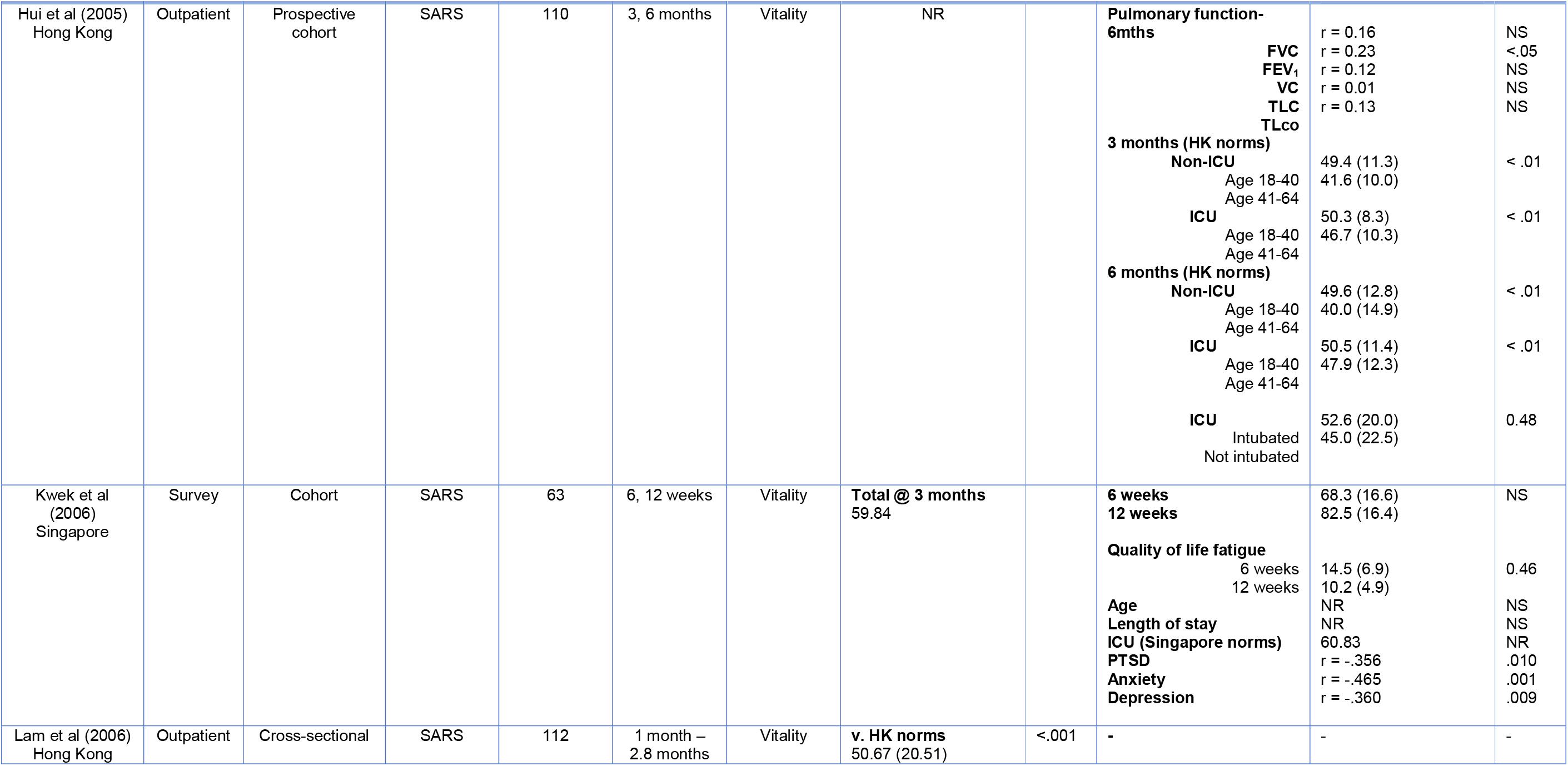
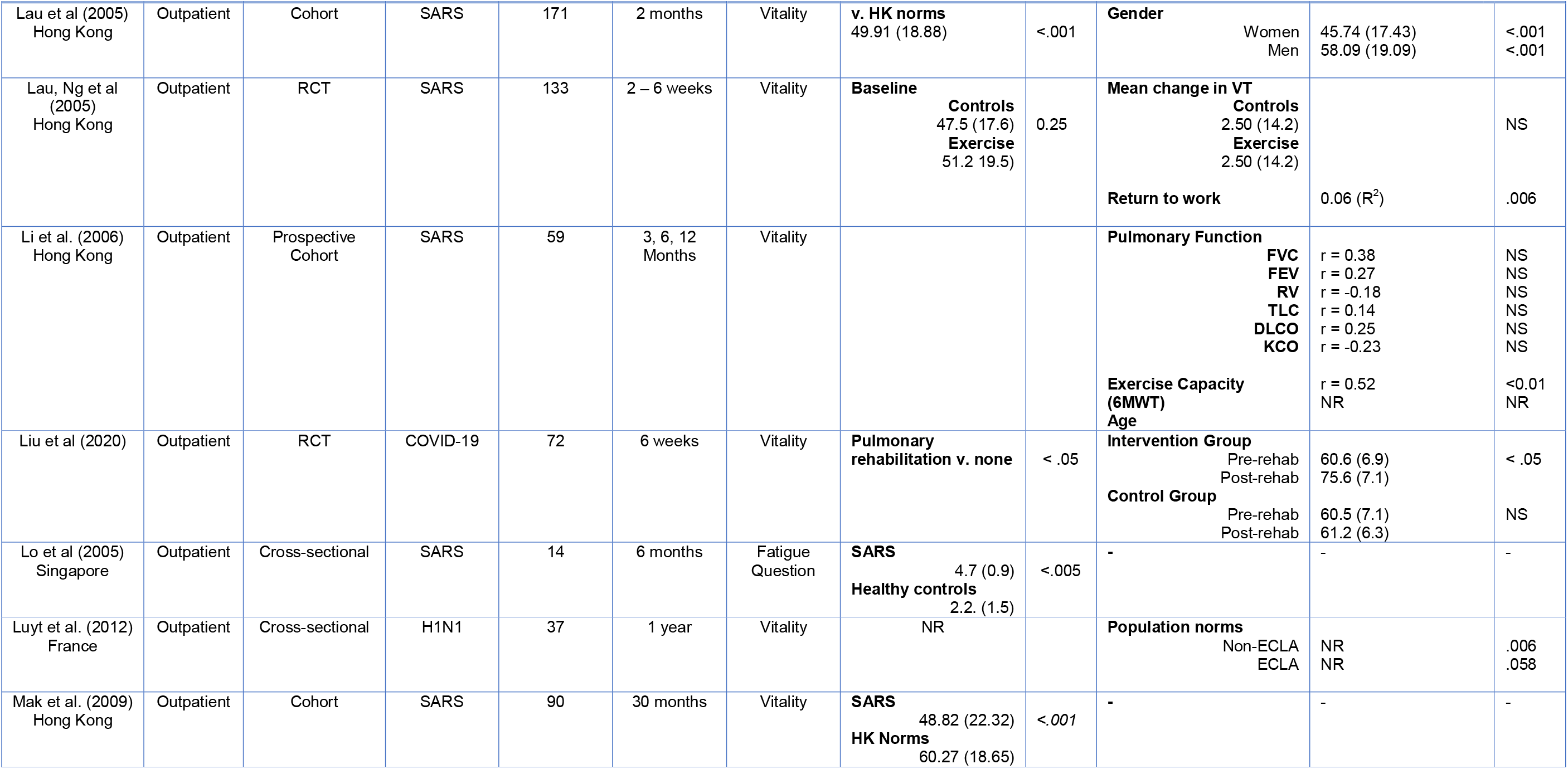
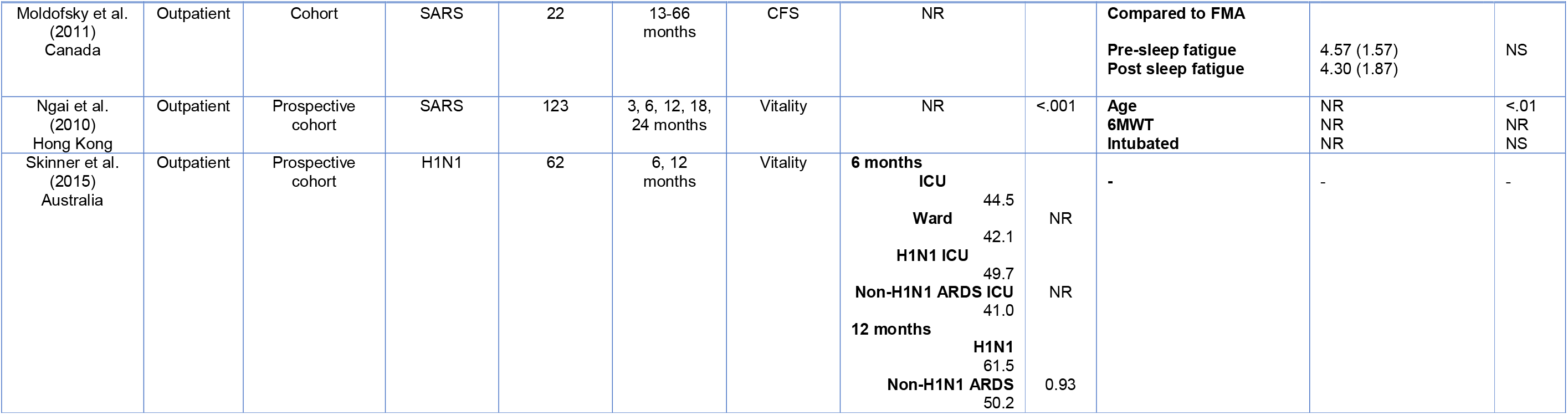
Summary of included studies

### Study characteristics

A total of 3,262 participants were represented in the included studies. There were 13 (40.6%) studies investigating fatigue/chronic fatigue using a fatigue scale and 19 (59.4%) studies using the ‘vitality’ subscale of the SF-36. The most common country of origin was Hong Kong with 10 studies, followed by China (7). Singapore, Canada and the UK (2 each). France, Saudi Arabia, Ireland, Korea, Australia and USA each had 1 study. SARS was the most frequently investigated virus (21 articles), COVID-19 (9), MERS (4) and H1N1 (3). One study investigated H7N9. Zero studies investigated fatigue succeeding the other named viruses. There were 15 prospective-cohort designs, 7 cross-sectional, 1 case-control, 5 cohort, 2 randomised-control and 3 systematic reviews. Time periods under study ranged from 2 weeks to 12 years post-discharge or infection (for community cases) with the most frequent time point being fatigue <3 months and 12 months post-discharge. For the purposes of the meta-analysis, time periods ranged from 2 weeks to 4 years (Appendix B). CASP quality assessments resulted in most studies receiving a ‘low’ or ‘moderate’ assessments indicated bias. Lower grades were assigned for multiple reasons including selection bias, lack of adequate control groups, small sample sizes and methodological bias (employment of unvalidated/unreliable scales).

### Factors associated with fatigue

Not all 32 studies investigated or reported factors associated with fatigue following a virus infection. Also, the available data for each risk factor were too few to conduct a quantified analysis, with a diverse number of outcome measures. However, associations that were available are illustrated in Table 2. Results are stratified by study design and univariate or multivariate results. Only available data was included in the table. In summary, four studies reported gender differences and depression. ICU admission was reported in 8 studies and 6 studies reported age/fatigue relationships. Anxiety, PTSD and exercise capacity in three studies. Two studies investigated pulmonary functions, length of stay, psychiatric morbidity, stigma and shorter response to follow-up. The remainder of risk factors were analysed by 1 study each.

**Table 2.**
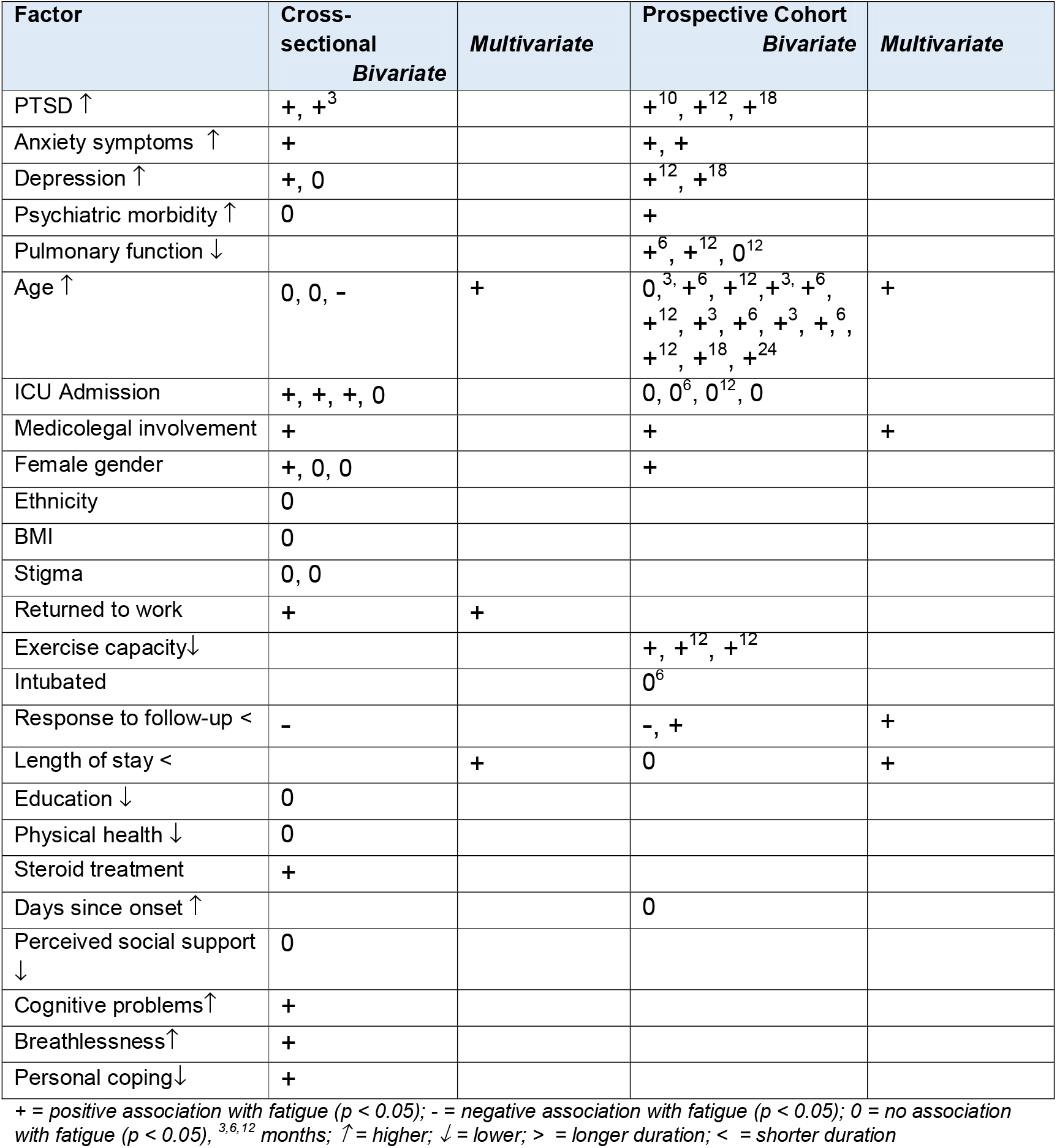
Variables associated with fatigue

| Factor | Cross-sectional<br>*Bivariate* | *Multivariate* | Prospective Cohort<br>*Bivariate* | *Multivariate* |
| --- | --- | --- | --- | --- |
| PTSD ↑ | +, +<sup>3</sup> |  | +<sup>10</sup>, +<sup>12</sup>, +<sup>18</sup> |  |
| Anxiety symptoms ↑ | + |  | +, + |  |
| Depression ↑ | +, 0 |  | +<sup>12</sup>, +<sup>18</sup> |  |
| Psychiatric morbidity ↑ | 0 |  | + |  |
| Pulmonary function ↓ |  |  | +<sup>6</sup>, +<sup>12</sup>, 0<sup>12</sup> |  |
| Age ↑ | 0, 0, - | + | 0,<sup>3</sup>, +<sup>6</sup>, +<sup>12</sup>, +<sup>3</sup>, +<sup>6</sup>,<br>+<sup>12</sup>, +<sup>3</sup>, +<sup>6</sup>, +<sup>3</sup>, +<sup>6</sup>,<br>+<sup>12</sup>, +<sup>18</sup>, +<sup>24</sup> | + |
| ICU Admission | +, +, +, 0 |  | 0, 0<sup>6</sup>, 0<sup>12</sup>, 0 |  |
| Medicolegal involvement | + |  | + | + |
| Female gender | +, 0, 0 |  | + |  |
| Ethnicity | 0 |  |  |  |
| BMI | 0 |  |  |  |
| Stigma | 0, 0 |  |  |  |
| Returned to work | + | + |  |  |
| Exercise capacity ↓ |  |  | +, +<sup>12</sup>, +<sup>12</sup> |  |
| Intubated |  |  | 0<sup>6</sup> |  |
| Response to follow-up < | - |  | -, + | + |
| Length of stay < |  | + | 0 | + |
| Education ↓ | 0 |  |  |  |
| Physical health ↓ | 0 |  |  |  |
| Steroid treatment | + |  |  |  |
| Days since onset ↑ |  |  | 0 |  |
| Perceived social support ↓ | 0 |  |  |  |
| Cognitive problems ↑ | + |  |  |  |
| Breathlessness ↑ | + |  |  |  |
| Personal coping ↓ | + |  |  |  |

+ = positive association with fatigue ( $p < 0.05$ ); - = negative association with fatigue ( $p < 0.05$ ); 0 = no association with fatigue ( $p < 0.05$ ), <sup>3,6,12</sup> months; ↑ = higher; ↓ = lower; > = longer duration; < = shorter duration

### Meta-analyses

Ten studies with a total of 1,310 participants were included for the proportional analysis using a random-effects model. A pooled prevalence from 10 studies was found to be 41% (95% CI 0.31-0.52). I^2^ statistic was 93% indicating ‘considerable’ heterogeneity. Details of this analysis are represented by a forest plot (Figure 2). Five studies reported means and standard deviations on SF-36 ‘vitality’ outcomes and were included for the means differences analysis. These results indicate vitality scores were −1.52 points lower for the participants compared to population norms (95% CI 13.53-10.49). This was non-significant p = 0.803. A forest plot of the results is available in Figure 3.

**Figure 2.**
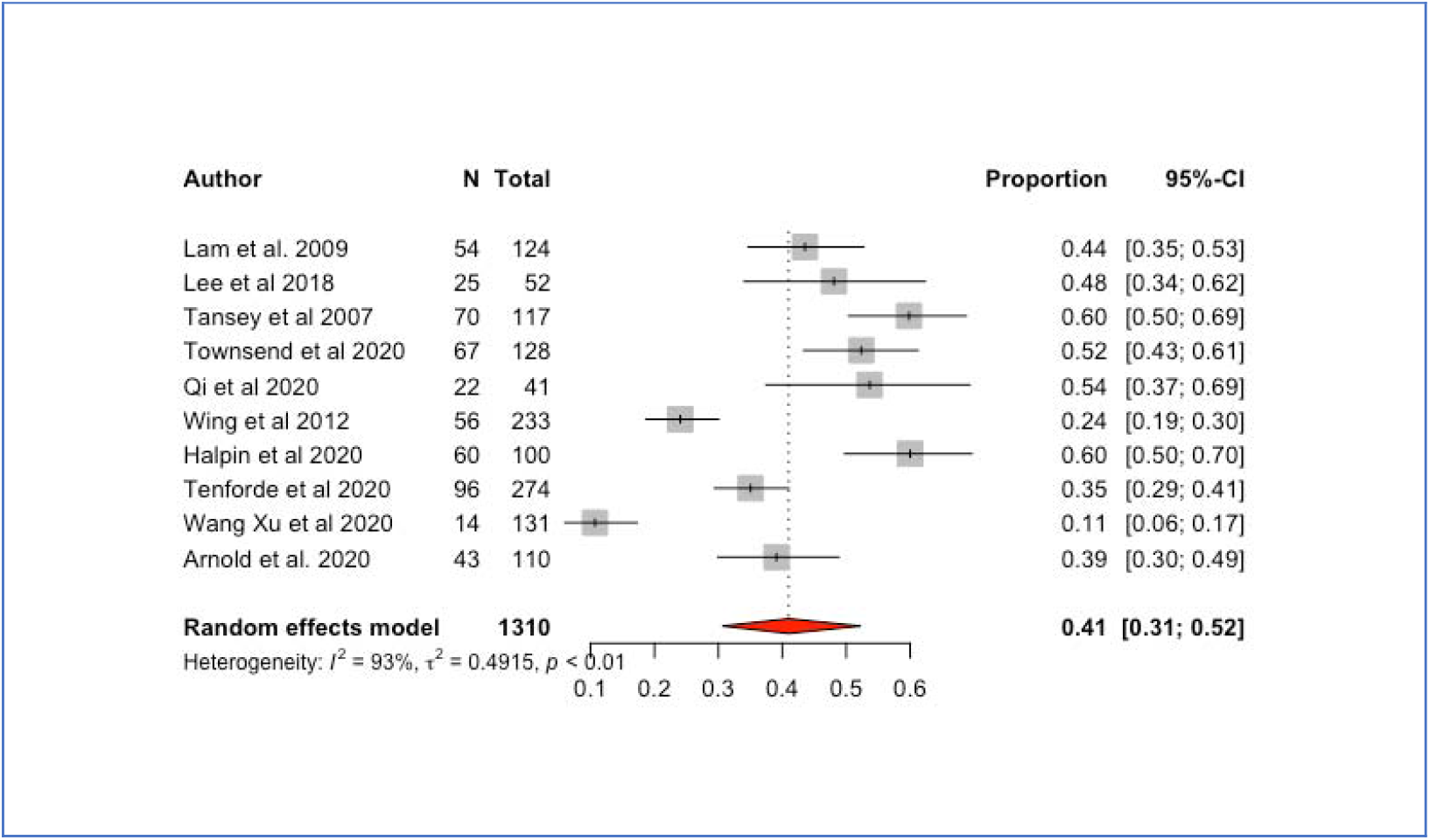
Forest plot for pooled prevalence for fatigue

**Figure 3.**
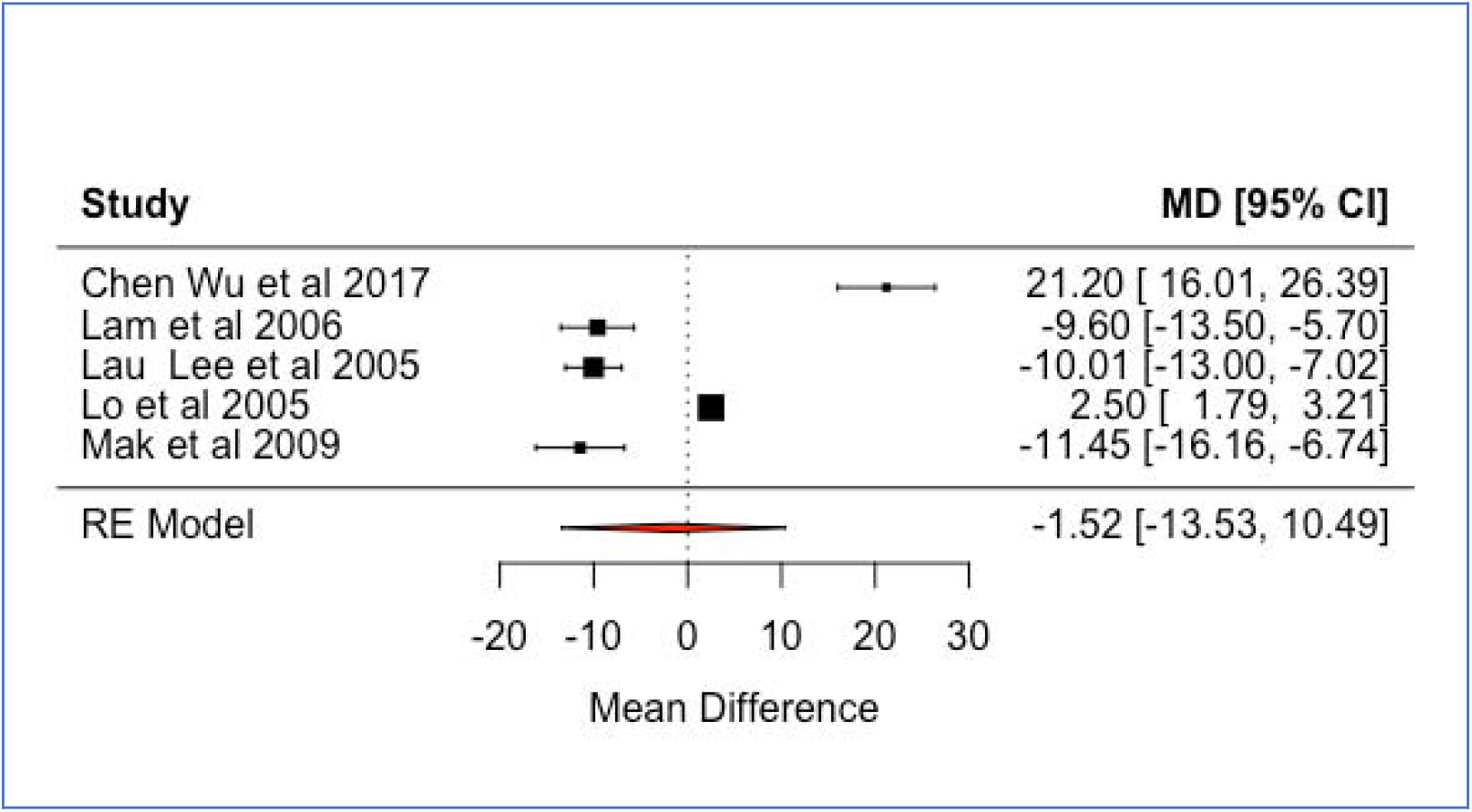
Forest plot for mean differences in vitality between survivors and population norms

## DISCUSSION

This review investigated the long-term fatigue outcomes in populations who had a confirmed coronavirus or other named virus diagnosis. We found 32 studies that provided data on fatigue and chronic fatigue, CFS and ‘vitality’. The majority of investigated viruses were SARS and COVID-19. MERS, H1N1 and 1 study examining H7N9 (‘avian’) made up the balance. No fatigue data was found among the remaining viruses. We found the pooled prevalence of fatigue to be considerable at 41% (95% CI 0.31-0.52) among 1,310 survivors and this fatigue was present between < 1 month (Tenforde et al. 2020; Wang et al., 2020) to 4 years post-infection (Lam et al., 2009). Our results are higher than one other study, which found a pooled estimate of 19.3% for fatigue in 316 survivors of SARS, MERS or COVID-19 (Rogers et al., 2020). To our knowledge no other studies have investigated fatigue systematically. The vitality subscale of the SF-36 (Ware & Sherborne, 1992) was used as a proxy for fatigue, with lower vitality scores representing higher fatigue. Our results indicated an overall lower mean for survivors compared to population norms. Although the results were not significant (p = 0.803), they suggest impaired functioning up to 2.5 years following exposure (Mak et al., 2009). Our results compare to previous systematic reviews examining vitality in recovered patients, which found lower mean vitality in SARS and MERS survivors compared to normative values (Ahmed et al., 2020). Similarly, in a literature review by Gardner et al. (2015), all domains of health-related quality of life measures, including vitality, were found to be lower than local population norms.

Fatigue and vitality changes over time were mixed, with some studies indicating improved scores between time points (Hong et al. 2009; Kwek et al. 2006; Lee et al. 2019) and other studies reporting no differences in fatigue for each follow-up period (Hui et al. 2005, Hui et al. 2005b). Nevertheless, most reported lower vitality or higher fatigue compared to population norms at all time points, up to 4 years post-infection (Lam et al. 2009; Li et al. 2006). We were not able to quantify the differences in fatigue between those who were admitted to ICU compared to ward patients, although fatigue for ICU groups was generally found to be higher compared to population norms (Hui et al. 2005; Hui et al. 2005b). Where studies compared ICU with ward groups, results indicated that fatigue was not significantly higher in the ICU group (Kwek et al., 2006; Li et al., 2006; Ngai et al., 2010; Skinner et al., 2015). Conversely, a recent study investigating COVID-19, found ‘new fatigue’ to be higher in ICU patients (72%) compared to the ward group, of whom 60.3% experienced fatigue (Halpin et al., 2020). In another study of COVID-19 patients (n. 110), Arnold et al., (2020) found worse fatigue among those who had more severe disease while MERS ICU groups had higher fatigue compared to non-ICU patients (p = 0.03) (Batawi et al. 2019).

Nor did our study calculate risk factors for fatigue due to the small number of studies investigating potential correlates, diverse outcome measures and aggregated SF-36 scores. Associations between fatigue and particular lung functions, however, were reported in 2 studies. For example, FVC and VC were found to be positively correlated with vitality at 3, 6 and 12 months (Hui et al. 2005) and FEV_1_ with higher vitality at 6 months (Hui et al. 2005b). Both suggest lower fatigue is associated with better pulmonary performance. Contrary evidence from an Australian H1N1 study, found that lung functions and fatigue for 62 survivors had improved to within normal range at 1-year post-discharge (Skinner et al., 2015). Exercise capacity, measured by the 6MWT, was lower than norms in most studies and correlated with fatigue in 3 studies (Hui et al., 2005; Li et al., 2006; Ngai et al., 2010). A systematic review found that mean walking distance was lower for all survivors compared to norms (Ahmed et al., 2020) with a ‘levelling’ of test results at 12 and 18 months (Ngai et al., 2010). Such limitations resulting from critical illness hospitalisation, are thought to contribute to reduced quality of life and fatigue outcomes in recovered patients (Herridge et al., 2003). However, these effects were observed independent of physical functioning. For example, in a study of 9 H1N1 recovered patients given pulmonary rehabilitation, there was a significant decrease in quality of life between 3 to 6 months, despite improved lung capacities and exercise capacity (Hsieh et al., 2018). In a study examining 1-year outcomes of SARS patients, Tansey et al. (2007) found that, while lung functions were within normal ranges, many reported persistent fatigue and shortness of breath.

Factors such as gender in COVID-19 (Chen et al., 2020; Lau et al., 2005; Qi et al., 2020; Townsend et al., 2020) and older age were found to be associated with fatigue (Chen et al., 2020; Hui et al., 2005; Hui et al., 2005b; Li et al., 2006; Ngai et al., 2010), although Halpin et al. (2020) found lower age was associated with higher fatigue in the ward group, while there were no differences between age groups at 3 months in Hui et al., (2005b). A multiplicity of social and physical risk factors for fatigue outcomes were investigated. For example, lower perceived social support in 41 COVID-19 patients was related to higher fatigue (Qi et al., 2020) and fatigue was reported as one of the reasons for not returning to work 12 years post-discharge (Guo et al., 2019), conversely predicting higher vitality among patients who had returned to work (Lau et al., 2005). Shorter duration of response to follow-up (Batawi et al., 2019; Lam et al., 2009), medicolegal involvement (Lam et al., 2009; Wing & Leung, 2012) and length of stay (Chen et al., 2020) were associated with higher fatigue. However, coping, health comorbidity and stigma were not found to be associated with fatigue (Qi et al., 2020).

Psychological factors were found to be significant factors for SARS and MERS survivors in systematic reviews (Gardner & Moallef, 2015; Rogers et al., 2020). Ahmed et al. (2020) found a pooled estimate for PTSD, depression and anxiety among SARS and MERS survivors to be 39%, 33% and 30% respectively. Among the included studies in our study chronic fatigue had a ‘reciprocal’ association with psychiatric diagnosis (Wing & Leung, 2012), while lower vitality was associated with PTSD symptomology (Halpin et al., 2020; Hong et al., 2009; Kwek et al., 2006), anxiety and depression (Kwek et al., 2006). Recovered MERS patients with chronic fatigue 1-year post exposure, had persistent PTSD symptoms mediated via depression 18 months post-infection (Lee et al., 2019). Moreover, those with psychiatric diagnosis were more likely to experience chronic fatigue 1 year later (Lam et al., 2009). In contrast, Qi et al., (2020) found no association between psychiatric morbidity and chronic fatigue in recovered COVID-19 patients.

Our study suggests that survivors of a serious influenza virus experienced considerable and long-lasting distress, physical impairments and psychological effects, including fatigue. The persistence of physical and psychological symptoms has been identified as a post-SARS sickness syndrome (Chrousos & Kaltsas, 2005). A syndrome distinguished by fatigue, myalgia, general weakness, psychological distress and sleep disorders. In respect of the current epidemic, there is potential for longer-term fatigue outcomes following a COVID-19 infection and current evidence points to a syndrome-like collection of symptoms. However, there are few studies examining probable correlates between fatigue or vitality outcomes with physical and pulmonary functioning; psychological or social functioning in recovering patients. Future studies should investigate such risk factors to help inform clinical interventions to address ‘long COVID’ symptomology. The generalisability of our results should be applied with caution due to the diversity of measurements, distance between investigation and exposure (i.e. < 1 month or > 2 years), variability in populations, different admission and discharge protocols and lung function reference ranges between countries (Chan, 2005).

## Data Availability

Data will be available after the review has been completed.

## APPENDIX A

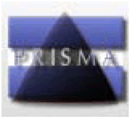 **PRISMA 2009 Flow Diagram**

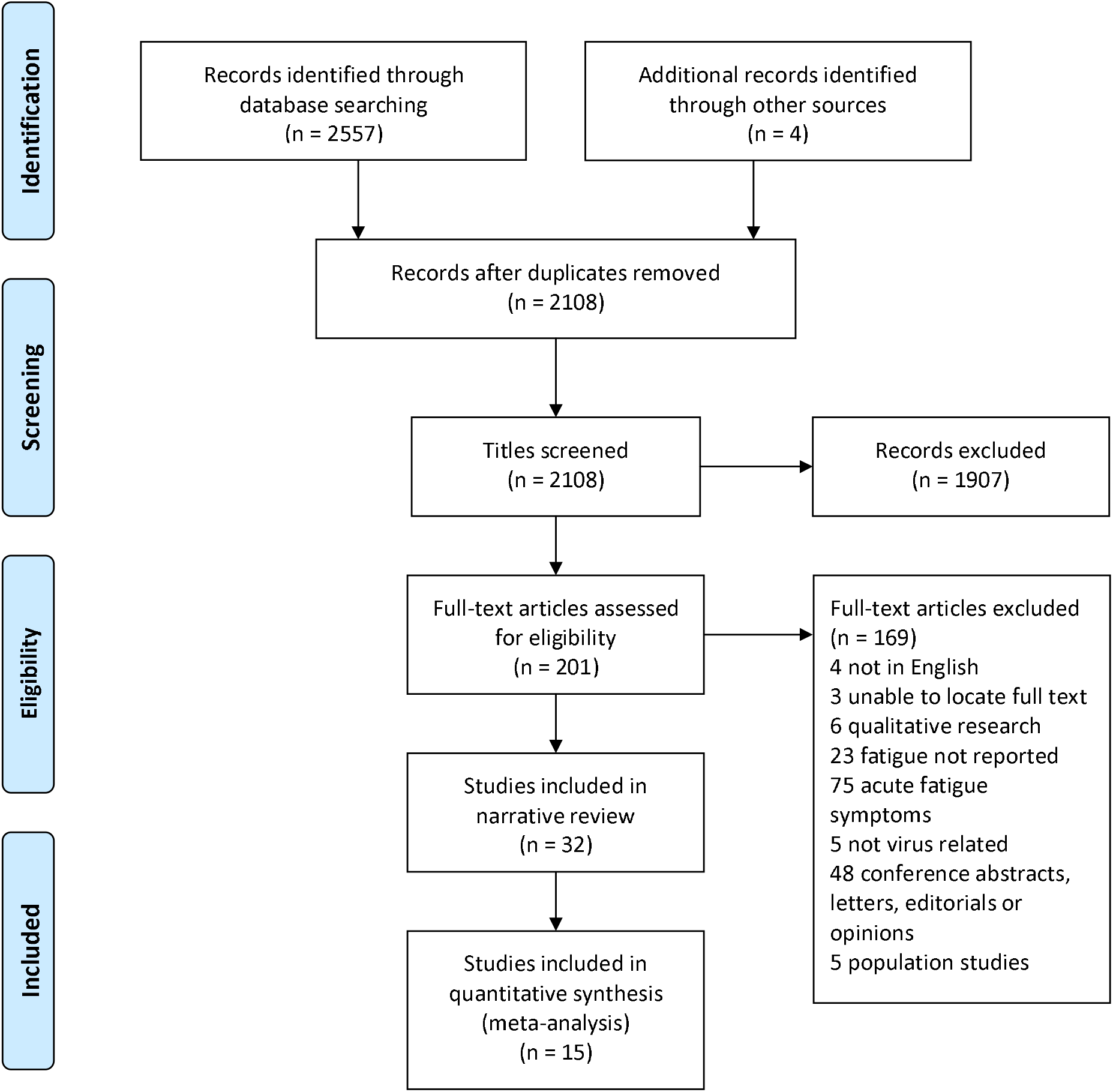

## APPENDIX B

*Studies included in pooled prevalence meta-analysis*

| **Study** | **Virus Type** | **Scale** | **Post-discharge** |
| --- | --- | --- | --- |
| **Lam et al. 2009** | SARS | Chalder Fatigue Scale | 4 years |
| **Lee et al. 2019** | MERS | Fatigue Severity Scale | 12 months |
| **Tansey et al. 2007** | SARS | Fatigue question | 12 months |
| **Townsend et al. 2020** | COVID-19 | Chalder Fatigue Scale | 2.5 months |
| **Qi et al. 2020** | COVID-19 | Chalder Fatigue Scale | 1 month |
| **Wing et al. 2012** | SARS | Fatigue question | 3 years |
| **Halpin et al. 2020** | COVID-19 | Interview question | 1 month |
| **Tenforde et al. 2020** | COVID-19 | Fatigue questionnaire | < 1 month |
| **Wang, Xu et. al. 2020** | COVID-19 | Fatigue question | < 1 month |
| **Arnold et al. 2020** | COVID-19 | Fatigue question | 3 months |

*Studies included in means differences meta-analysis*

*Studies included in means differences meta- analysis*
| **Study** | **Virus Type** | **Scale** | **Post-discharge** |
| --- | --- | --- | --- |
| **Chen, Wu et al. 2017** | H7N9 | SF-36 | 3 months |
| **Lam et al. 2006** | SARS | SF-36 | < 3 months |
| **Lau, Lee et al. 2005** | SARS | SF-36 | < 3 months |
| **Lo et al. 2005** | SARS | ADQ | 6 months |
| **Mak et al. 2009** | SARS | SF-36 | 30 months |

